# Characteristics and outcomes of 7620 Multiple Sclerosis patients admitted with COVID-19 in the United States

**DOI:** 10.1101/2023.02.15.23285994

**Authors:** Kamleshun Ramphul, Shaheen Sombans, Renuka Verma, Petras Lohana, Balkiranjit Kaur Dhillon, Stephanie Gonzalez Mejias, Sailaja Sanikommu, Yogeshwaree Ramphul, Prince Kwabla Pekyi-Boateng

## Abstract

**Background:** At the start of the COVID-19 pandemic, several experts raised concerns about its impact on Multiple Sclerosis (MS) patients. Several small sample studies were published throughout the pandemic highlighting certain risk factors and outcomes. This study aims to provide a perspective using the biggest inpatient database from the United States.

**Method:** We screened for COVID-19 cases between April to December 2020, via the 2020 National Inpatient Sample (NIS). Characteristics of COVID-19 patients with and without MS were studied. The odds of mortality, mechanical ventilation and non-invasive ventilation were also analyzed. Finally, we investigated the risk factors of various outcomes among MS patients.

**Results:** We identified 1,628,110 hospitalizations with COVID-19, including 7620 (0.5%) MS patients. 68.6% of MS cases were Whites, and 63.3% were covered by Medicare. Compared to non-MS patients, MS patients with COVID-19 were mostly Females, had depression, peripheral vascular disease, and smoked. However, MS patients had lower cases of alcohol abuse, obesity, hyperlipidemia, diabetes, hypertension, CKD, or maintenance dialysis. MS patients with COVID-19 were also younger (mean age 60.65 years vs. 62.60 years, p<0.01). 8.9% of MS patients with COVID-19 did not survive their hospitalization, and it was lower than non-MS cases (12.9%, aOR 0.783, 95% CI 0.721-0.852, p<0.01). Less MS patients with COVID-19 needed non-invasive ventilation (4.5% vs. 6.4%, aOR 0.790, 95% CI 0.706-0.883, p<0.01) and mechanical ventilation (9.0% vs. 11.2%, aOR 1.017, 95% CI 0.937-1.104, p=0.687).

Furthermore, MS patients with COVID-19 reported higher odds of non-invasive ventilation if they were of ages 60 and above (aOR 2.124, p<0.01), had chronic pulmonary disease (aOR 1.691, p<0.01), obesity (aOR 1.69, p<0.01), and diabetes (aOR 1.573, p<0.01). Private insurance beneficiaries showed reduced risk compared to Medicare (aOR 0.523, p<0.01). Similarly, for mechanical ventilation, those ages 60 and above (aOR 1.404, p<0.01), alcohol abuse (aOR 6.404, p<0.01), obesity (aOR 1.417, p<0.01), diabetes (aOR 1.992, p<0.01), hypertension (aOR 1.269, p=0.016), or dialysis (aOR 3.003, p<0.01) had higher odds, while females (aOR 0.700, p<0.01), smokers (aOR 0.588, p<0.01), and those with depression (aOR 0.698, p<0.01) or hyperlipidemia (aOR 0.711, p<0.01) showed reduced odds.

Our study further found higher odds of mortality among those of age 60 and above (aOR 3.813, p<0.01), chronic pulmonary disease (aOR 1.739, p<0.01), obesity (aOR 1.425, p<0.01), CKD (aOR 1.982, p<0.01), or a history of old MI (aOR 1.864, p<0.01) while females (aOR 0.610, p<0.01), smokers (aOR 0.770, p<0.01), as well as those with depression (aOR 0.695, p<0.01), and hyperlipidemia (aOR 0.769, p<0.01) showed better outcomes. Blacks had lower odds of dying (aOR 0.636, p<0.01), whereas Hispanics had higher odds of dying (aOR 1.674, p<0.01), compared to Whites. Medicaid and Privately insured patients had lower odds of dying compared to Medicare i.e. (aOR 0.435, p<0.01), and (aOR 0.488, p<0.01), respectively.

**Conclusion:** We found several differences in patient characteristics among MS and non-MS patients with COVID-19. MS patients were also less likely to die or require non-invasive ventilation than non-MS patients. Further risk factors influencing the different outcomes among MS patients were also identified.

## Introduction

Multiple sclerosis (MS), an autoimmune disease, can cause various inflammatory insults, leading to motor impairment and other clinical findings and disabilities. The pandemic of coronavirus disease 2019 (COVID-19) has infected over 650 million people across the world over its three years, leading to six million deaths.(1) The elderly and those with multiple comorbidities or immunosuppressed state often experience a more severe course of the disease and poorer prognosis.(2) Since MS patients have an increased risk of infection in general, and several patients are also on disease-modifying therapies (DMTs), some studies hypothesized that MS patients might also be at risk for a more severe outcome with the pandemic.(3)

Early results from different studies have been conflicting, mostly due to the small sample size. We, therefore, proceeded to conduct a large retrospective analysis, via the most extensive inpatient database in the US to provide a more accurate estimate.

## Method

Each year, the Healthcare Cost and Utilization Project (HCUP), in association with the Agency for Healthcare Research and Quality’s (AHRQ) and their partners, develop and release the National Inpatient Sample (NIS), which can cover around 35 million hospitalization records in its weighted form. The 2020 NIS, which is the most recent version released, was used for our study.(4)

We identified patients with a diagnosis of COVID-19 using the ICD-10 code “U071” which has shown high sensitivity and specificity in past studies.(5) As the code was introduced on April 1^st^, 2020, we improved the accuracy of our study by filtering for patients admitted only between April 1^st^, 2020 and December 31^st^, 2020. A diagnosis of Multiple Sclerosis was also identified via the ICD-10 code “G35” as in past recommendations.(6, 7) Several different comorbidities and patient characteristics were also found via the appropriate codes from HCUP and previous studies.(8-11)

Our analysis was done in three steps. First, we used Chi-square tests to view and compare patient characteristics between MS and non-MS cases that were positive for COVID-19. Then the adjusted odds ratio (aOR) of MS patients requiring mechanical ventilation, non-invasive ventilation, or not surviving their hospitalization was assessed via logistic regression models, compared to non-MS patients. Finally, we isolated all cases of MS patients with COVID-19 to evaluate for predictors of mechanical ventilation, non-invasive ventilation, and mortality among them.

### Ethical clearance

HCUP provides the NIS in a de-identified form and exempts users from requiring IRB approvals. Moreover, the DUA from the organization also waives the need for ethical approval.

## Results

Our study found 1,628,110 cases of COVID-19 among patients admitted between April 1^st^, 2020 to December 31^st^, 2020 in the United States. We further found that 7620 (0.5%) had a diagnosis of Multiple Sclerosis (table 1). Among all the patients admitted with COVID-19, those with MS were more likely to be female (65.7% vs. 48.1%, p<0.01), report a diagnosis of depression (25.3% vs. 10.9%, p<0.01), peripheral vascular disease (4.9% vs. 4.0%, p<0.01) or be a smoker (nicotine/tobacco use) (29.6% vs. 25.7%, p<0.01). Racial disparities were also noticed, as a higher percentage of MS patients were White (68.6%, p<0.010). Medicare was the preferred insurance form covering MS patients (63.3%, p<0.01). MS patients also reported a lower incidence of various conditions such as alcohol abuse (0.9% vs. 2.3%, p<0.01), obesity (24.1 vs. 26.2%, p<0.01), hyperlipidemia (35.4 vs.39.5, p<0.01), diabetes (28.7 vs.40.0%, p<0.01), hypertension (58.1% vs. 64.0%, p<0.01), CKD (13.8% vs. 21.4%, p<0.01), maintenance dialysis (1.0% vs. 3.2%, p<0.01) and fewer patients were of ages 60 or more (56.7%, vs. 61.4%, p<0.01) (mean age 60.65 years vs. 62.60 years, p<0.01).

**Table 1.** Characteristics of COVID-19 positive patients with or without a history of Multiple Sclerosis, hospitalized in the United States between April to December 2020.

| Characteristics | Multiple Sclerosis<br>(n=7620) (%) | Non- Multiple<br>Sclerosis patients<br>(n=1620490) (%) | p-value |
| --- | --- | --- | --- |
| Patient Characteristics |  |  |  |
| Female | 65.7 | 48.1 | <0.01 |
| Age<br>60 and more | 56.7 | 61.4 | <0.01 |
| Race |  |  | <0.01 |
| White | 68.6 | 51.0 |  |
| Black | 21.0 | 18.7 |  |
| Hispanic | 7.5 | 21.8 |  |
| Others | 2.8 | 8.5 |  |
| Insurance |  |  | <0.01 |
| Medicare | 63.3 | 50.2 |  |
| Medicaid | 10.2 | 15.1 |  |
| Private | 23.3 | 26.2 |  |
| Other forms | 3.2 | 8.6 |  |
| Comorbidities |  |  |  |
| Autoimmune conditions | 3.5 | 3.1 | 0.084 |
| Alcohol abuse | 0.9 | 2.3 | <0.01 |
| Depression | 25.3 | 10.9 | <0.01 |
| Drug abuse | 1.8 | 2.0 | 0.394 |
| Chronic pulmonary disease | 21.5 | 21.5 | 0.966 |
| Obesity | 24.1 | 26.2 | <0.01 |
| Peripheral vascular disease | 4.9 | 4.0 | <0.01 |
| Hyperlipidemia | 35.4 | 39.5 | <0.01 |
| Nicotine or tobacco use | 29.6 | 25.7 | <0.01 |
| Diabetes | 28.7 | 40.0 | <0.01 |
| Hypertension | 58.1 | 64.0 | <0.01 |
| Old myocardial infarction | 3.8 | 4.1 | 0.145 |
| CKD | 13.8 | 21.4 | <0.01 |
| Maintenance Dialysis | 1.0 | 3.2 | <0.01 |
| Outcomes |  |  |  |
| Died | 8.9 | 12.9 | <0.01 |
| Mechanical ventilation | 9.0 | 11.2 | <0.01 |
| Non-invasive ventilation | 4.5 | 6.4 | <0.01 |

A lower mortality rate was observed among COVID-19 positive MS patients compared to those with COVID-19 and not having a diagnosis of MS (8.9% vs. 12.9%, aOR 0.783, 95% CI 0.721-0.852, p<0.01). Moreover, MS patients were also less likely to require non-invasive ventilation (4.5% vs. 6.4%, aOR 0.790, 95% CI 0.706-0.883, p<0.01). While only 9.0% of MS cases needed mechanical ventilation (vs. 11.2% among non-MS patients), after adjusting for variables, no statistical significance was found (aOR 1.017, 95% CI 0.937-1.104, p=0.687).

Finally, our study found several potential predictors among MS patients that influenced their odds of requiring non-invasive ventilation, mechanical ventilation, or mortality.

### Non-invasive ventilation

The odds of requiring non-invasive ventilation were higher in patients aged 60 and above (aOR 2.124, 95% CI 1.574-2.867, p<0.01), as well as those with chronic pulmonary disease (aOR 1.691, 95% CI 1.31-2.184, p<0.01), obesity (aOR 1.69, 95% CI 1.316-2.17, p<0.01), and diabetes (aOR 1.573, 95% CI 1.229-2.013, p<0.01). Meanwhile, it was also noted that patients covered by private insurance showed lower odds of requiring non-invasive ventilation compared to Medicare beneficiaries (aOR 0.523, 95% CI 0.364-0.751, p<0.01).

### Mechanical ventilation

MS patients with COVID-19 were more likely to require mechanical ventilation if they were aged 60 and above (aOR 1.404, 95% CI 1.149-1.717, p<0.01), with underlying alcohol abuse (aOR 6.404, 95% CI 3.373-12.160, p<0.01), obesity (aOR 1.417, 95% CI 1.172-1.713, p<0.01), diabetes (aOR 1.992, 95% CI 1.663-2.386, p<0.01), hypertension (aOR 1.269, 95% CI 1.046-1.539, p=0.016), or maintenance dialysis (aOR 3.003, 95% CI 1.682-5.363, p<0.01). However, the odds of requiring mechanical ventilation were noted to be lower in females (aOR 0.700, 95% CI 0.588-0.833, p<0.01), nicotine/tobacco users (aOR 0.588, 95% CI 0.477-0.725, p<0.01), and those diagnosed with depression (aOR 0.698, 95% CI 0.562-0.866, p<0.01) or hyperlipidemia (aOR 0.711, 95% CI 0.587-0.860, p<0.01).

### Mortality

Finally, we found that MS patients of age 60 and above (aOR 3.813, 95% CI 2.957-4.917, p<0.01), or those with chronic pulmonary disease (aOR 1.739, 95% CI 1.429-2.117, p<0.01), obesity (aOR 1.425, 95% CI 1.167-1.741, p<0.01), CKD (aOR 1.982, 95% CI 1.598-2.458, p<0.01), or a history of old MI (aOR 1.864, 95% CI 1.320-2.633, p<0.01) had higher odds of inpatient death during their hospitalization. On the contrary, females (aOR 0.610, 95% CI 0.511-0.728, p<0.01), smokers (aOR 0.770, 95% CI 0.633-0.937, p<0.01), as well as those with depression (aOR 0.695, 95% CI 0.561-0.862, p<0.01), and hyperlipidemia (aOR 0.769, 95% CI 0.638-0.928, p<0.01) showed a better outcome. Racial differences were also observed as Blacks had lower odds of dying (aOR 0.636, 95% CI 0.493-0.819, p<0.01), whereas Hispanics had higher odds of dying (aOR 1.674, 95% CI 1.175-2.386, p<0.01), compared to Whites. Medicaid and Privately insured patients had lower odds of dying compared to Medicare i.e. (aOR 0.435, 95% CI 0.272-0.697, p<0.01), and (aOR 0.488, 95% CI 0.365-0.652, p<0.01) respectively.

## Discussion

Our study from the 2020 NIS provides one of the most extensive and up-to-date information on the impact of COVID-19 among patients who also have a diagnosis of Multiple Sclerosis. Several differences in patient characteristics with MS and positive for COVID-19 are similar to the overall distribution among patients with multiple sclerosis in the United States. A higher female-to-male ratio, a larger percentage of patients being classified as White, a more significant proportion of MS patients being Medicare beneficiaries, and age characterizations involving more patients being below 60 than above, all correspond to the findings previously reported from the 2019 NIS (6). This also reflects trends in various studies done in other countries (6, 12, 13) and runs parallel with the census distribution in the United States.(6)

Furthermore, the lower incidence of various comorbidities in general, among all MS patients with COVID-19 vs non-MS patients with COVID-19, such as diabetes, alcohol abuse, obesity, hyperlipidemia, hypertension, and maintenance dialysis may be linked with a higher proportion of MS patients with COVID-19 being younger and may be under regular medical care from a younger age, thus may have been advised to adhere to a healthier lifestyle. Our findings also confirmed that MS patients with COVID-19 had a lower risk of mortality and were less likely to require non-invasive ventilation. Similar results were reported in Iran, where the rate of mortality was lower among MS patients compared to the rest of the population.(14) MS patients also use different medications that may influence the impact of COVID-19. Unfortunately, the NIS does not allow an in-depth analysis of such medications, making it a possible limitation of our study.

Various patient characteristics and comorbidities also influenced outcomes among MS patients. In our study, the elderly (ages 60 and more) with MS were at higher risk for non-invasive ventilation, mechanical ventilation, and mortality. In their analysis, Prosperini et al. found similar results as the case-fatality rate was higher among older COVID-19-positive MS patients, with an “exponential increase above 60 years”.(15) Multiple age-related changes and weakening of the immune response in the elderly, as also seen in non-MS elderly patients, may be one of the primary reasons behind these findings.(16) Furthermore, we also linked chronic pulmonary disease, obesity, diabetes, alcohol abuse, hypertension, dialysis use, chronic kidney disease, and presence of an old MI to poorer outcomes among MS patients, as already reported in the general population.(17-25)

Finally, our study reported racial disparities as Blacks with MS admitted for COVID-19 had lower odds of death than Whites, while Hispanics showed increased risk. This contradicts past results from Salter et al. based on the North American Registry, whereby the mortality rate among Whites with MS and COVID-positive was estimated at 3.5%, while it was 4.2% among Blacks, and 1.1% among Hispanics. However, the sample size used in their study was smaller, and since there are also some limitations to our study, such as the inability to adjust for medications, further studies from direct hospital data may help in understanding these disparities.(26) Sex differences were also observed in our study, which conforms with other studies.(27) The impact of COVID-19 among smokers has been widely debated as some studies linked them with a lower risk of infection, while others reported the opposite.(28-32) Our study also found lower odds of mechanical ventilation and death among those with depression, however, this association is not fully understood. As the pandemic is believed to have caused psychological impacts on many individuals, leading to a rise in the number of anxiety and depressive episodes across the world(33), neurologists and primary care providers must screen MS patients, who already have a higher incidence of depression in general, for any warning signs to provide help in a timely manner.(34, 35)

Our analysis provided a significant perspective, via a big sample size, on the characteristics and outcomes of MS patients that were admitted with COVID-19. However, the NIS database has some limitations. As patient medications and management plans are not provided under NIS, these aspects could not be studied. In addition, their post-discharge follow-ups could not be tracked and the changes in outcomes and long-term prognosis and side-effects could also not be monitored. Future studies should target these shortcomings, which can be done in a clinical setting via a retrospective analysis of patient files and adequate follow-ups. However, our analysis managed to provide several conclusions that may help improve the care of MS patients during the pandemic and prepare neurologists for future pandemics.

## Conclusion

We studied the characteristics of MS patients admitted with COVID-19 in 2020, and identified various risk factors that can influence the need for non-invasive ventilation or mechanical ventilation, as well as risk factors influencing mortality.

## Data Availability

All data produced in the present study are available upon reasonable request to the authors

## Acknowledgement

We are thankful to HCUP, AHRQ, and partners for the database. https://www.hcup-us.ahrq.gov/db/hcupdatapartners.jsp.

## References

1. [Available from: https://www.worldometers.info/coronavirus/country/us/.

2. Wu Z, McGoogan JM. Characteristics of and Important Lessons From the Coronavirus Disease 2019 (COVID-19) Outbreak in China: Summary of a Report of 72L314 Cases From the Chinese Center for Disease Control and Prevention. Jama. 2020;323(13):1239–42.

3. Luna G, Alping P, Burman J, Fink K, Fogdell-Hahn A, Gunnarsson M, et al. Infection Risks Among Patients With Multiple Sclerosis Treated With Fingolimod, Natalizumab, Rituximab, and Injectable Therapies. JAMA neurology. 2020;77(2):184–91.

4. HCUP National Inpatient Sample (NIS). Healthcare Cost and Utilization Project (HCUP). 2020. Agency for Healthcare Research and Quality, Rockville, MD. 2022 [Available from: http://www.hcup-us.ahrq.gov/nisoverview.jsp.

5. Kadri SS, Gundrum J, Warner S, Cao Z, Babiker A, Klompas M, et al. Uptake and Accuracy of the Diagnosis Code for COVID-19 Among US Hospitalizations. Jama. 2020;324(24):2553–4.

6. Ramphul K, Lohana P, Verma R, Kumar N, Ramphul Y, Lohana A, et al. An epidemiological analysis of multiple sclerosis patients hospitalized in the United States. Multiple sclerosis and related disordersx. 2022;63:103840.

7. Oud L, Garza J. Association of multiple sclerosis with mortality in sepsis: a population-level analysis. Journal of intensive care. 2022;10(1):36.

8. Hirode G, Saab S, Wong RJ. Trends in the Burden of Chronic Liver Disease Among Hospitalized US Adults. JAMA network open. 2020;3(4):e201997.

9. Kichloo A, Jamal S, Albosta M, Khan MZ, Aljadah M, Edigin E, et al. Increased inpatient mortality in patients hospitalized for atrial fibrillation and atrial flutter with concomitant amyloidosis: Insight from National Inpatient Sample (NIS) 2016-2017. Indian pacing and electrophysiology journal. 2021;21(6):344–8.

10. Ramphul K, Kumar N, Verma R, Ramphul Y, Sombans S, Kumari K, et al. Acute myocardial infarction in patients with multiple sclerosis; An insight from 1785 cases in the United States. Multiple sclerosis and related disorders. 2022;68:104140.

11. Yang CW, Li S, Dong Y, Paliwal N, Wang Y. Epidemiology and the Impact of Acute Kidney Injury on Outcomes in Patients with Rhabdomyolysis. Journal of clinical medicine. 2021;10(9).

12. Türk Börü Ü, Duman A, Kulualp A, Güler N, Tasdemir M, Yilmaz Ü, et al. Multiple sclerosis prevalence study: The comparison of 3 coastal cities, located in the black sea and mediterranean regions of Turkey. Medicine. 2018;97(42):e12856.

13. Ahlgren C, Odén A, Lycke J. High nationwide prevalence of multiple sclerosis in Sweden. Multiple sclerosis (Houndmills, Basingstoke, England). 2011;17(8):901–8.

14. Ghadiri F, Sahraian MA, Shaygannejad V, Ashtari F, Ghalyanchi Langroodi H, Baghbanian SM, et al. Characteristics of COVID-19 in patients with multiple sclerosis. Multiple sclerosis and related disorders. 2022;57:103437.

15. Prosperini L, Tortorella C, Haggiag S, Ruggieri S, Galgani S, Gasperini C. Increased risk of death from COVID-19 in multiple sclerosis: a pooled analysis of observational studies. Journal of neurology. 2022;269(3):1114–20.

16. Bsteh G, Bitschnau C, Hegen H, Auer M, Di Pauli F, Rommer P, et al. Multiple sclerosis and COVID-19: How many are at risk? European journal of neurology. 2021;28(10):3369–74.

17. Gerayeli FV, Milne S, Cheung C, Li X, Yang CWT, Tam A, et al. COPD and the risk of poor outcomes in COVID-19: A systematic review and meta-analysis. EClinicalMedicine. 2021;33:100789.

18. Wang QQ, Kaelber DC, Xu R, Volkow ND. Correction: COVID-19 risk and outcomes in patients with substance use disorders: analyses from electronic health records in the United States. Molecular psychiatry. 2021;26(1):40.

19. Singh R, Rathore SS, Khan H, Karale S, Chawla Y, Iqbal K, et al. Association of Obesity With COVID-19 Severity and Mortality: An Updated Systemic Review, Meta-Analysis, and Meta-Regression. Frontiers in endocrinology. 2022;13:780872.

20. Ramphul K, Lohana P, Ramphul Y, Park Y, Mejias S, Dhillon BK, et al. Hypertension, diabetes mellitus, and cerebrovascular disease predispose to a more severe outcome of COVID-19. Archives of medical sciences Atherosclerotic diseases. 2021;6:e30–e9.

21. Kin KC, Zhou H, Gysi M, Huang CW, Selevan DC, Lu DD, et al. Outcomes Among Hospitalized Patients With COVID-19 and Acute Kidney Injury Requiring Renal Replacement Therapy. The Permanente journal. 2022;26(3):39–45.

22. Gok M, Cetinkaya H, Kandemir T, Karahan E, Tuncer İ B, Bukrek C, et al. Chronic kidney disease predicts poor outcomes of COVID-19 patients. International urology and nephrology. 2021;53(9):1891–8.

23. Drager LF, Pio-Abreu A, Lopes RD, Bortolotto LA. Is Hypertension a Real Risk Factor for Poor Prognosis in the COVID-19 Pandemic? Current hypertension reports. 2020;22(6):43.

24. Corona G, Pizzocaro A, Vena W, Rastrelli G, Semeraro F, Isidori AM, et al. Diabetes is most important cause for mortality in COVID-19 hospitalized patients: Systematic review and meta-analysis. Reviews in endocrine & metabolic disorders. 2021;22(2):275–96.

25. Bonow RO, Fonarow GC, O’Gara PT, Yancy CW. Association of Coronavirus Disease 2019 (COVID-19) With Myocardial Injury and Mortality. JAMA cardiology. 2020;5(7):751–3.

26. Salter A, Fox RJ, Newsome SD, Halper J, Li DKB, Kanellis P, et al. Outcomes and Risk Factors Associated With SARS-CoV-2 Infection in a North American Registry of Patients With Multiple Sclerosis. JAMA neurology. 2021;78(6):699–708.

27. Ramírez-Soto MC, Ortega-Cáceres G, Arroyo-Hernández H. Sex differences in COVID-19 fatality rate and risk of death: An analysis in 73 countries, 2020-2021. Le infezioni in medicina. 2021;29(3):402–7.

28. Paleiron N, Mayet A, Marbac V, Perisse A, Barazzutti H, Brocq FX, et al. Impact of Tobacco Smoking on the Risk of COVID-19: A Large Scale Retrospective Cohort Study. Nicotine & tobacco research : official journal of the Society for Research on Nicotine and Tobacco. 2021;23(8):1398–404.

29. Reddy RK, Charles WN, Sklavounos A, Dutt A, Seed PT, Khajuria A. The effect of smoking on COVID-19 severity: A systematic review and meta-analysis. Journal of medical virology. 2021;93(2):1045–56.

30. Lombardi C, Roca E, Ventura L, Cottini M. Smoking and COVID-19, the paradox to discover: An Italian retrospective, observational study in hospitalized and non-hospitalized patients. Medical hypotheses. 2021;146:110391.

31. Guo FR. Active smoking is associated with severity of coronavirus disease 2019 (COVID-19): An update of a meta-analysis. Tobacco induced diseases. 2020;18:37.

32. Xie J, Zhong R, Wang W, Chen O, Zou Y. COVID-19 and Smoking: What Evidence Needs Our Attention? Frontiers in physiology. 2021;12:603850.

33. Moayed MS, Vahedian-Azimi A, Mirmomeni G, Rahimi-Bashar F, Goharimoghadam K, Pourhoseingholi MA, et al. Depression, Anxiety, and Stress Among Patients with COVID-19: A Cross-Sectional Study. Advances in experimental medicine and biology. 2021;1321:229–36.

34. Siegert RJ, Abernethy DA. Depression in multiple sclerosis: a review. Journal of neurology, neurosurgery, and psychiatry. 2005;76(4):469–75.

35. Global prevalence and burden of depressive and anxiety disorders in 204 countries and territories in 2020 due to the COVID-19 pandemic. Lancet (London, England). 2021;398(10312):1700–12.

